# Assessing the Economic Burden of Alcohol Use in India Compared to Excise Taxes on Alcohol Sales

**DOI:** 10.1101/2023.09.05.23295088

**Authors:** Siddhesh Zadey

## Abstract

**Background:** Alcohol use is among the topmost public health risk factors. There is a dearth of evidence on the broader impact of alcohol use on society, especially, for lower-middle-income countries like India. We retrospectively studied the economic burden of alcohol use and compared it with revenue from excise taxes on alcohol sales for India and its states in 2019.

**Methods:** For estimating the overall alcohol-attributable disease burden, disability-adjusted life-years or DALYs for India and its 30 states were extracted for all causes, all age groups, and both sexes with alcohol use as a risk factor from the Global Burden of Disease 2019 Study. Economic burden was calculated by the value of life-year (VLY) or full-income approach. National and state-level (top 10 revenue-earning states) data on excise taxes on alcohol sales were acquired from the Reserve Bank of India. Net losses were assessed by subtracting the revenue from the economic burden. All values are in Indian rupees (INR). Sensitivity analysis was conducted using different discount rates (0%, 3%, and 7%) for VLY values while uncertainty was propagated using 95% uncertainty intervals (UIs) for DALYs.

**Results:** The economic burden of alcohol use in India was 6.2 (95% UI: 4.8, 7.8) trillion INR in 2019. State-level burden varied from 7.3 (4.7, 10.4) billion INR in Mizoram to 1.1 (0.8, 1.4) trillion INR in Maharashtra. Nationally, revenue was 1.8 trillion INR depicting a net loss of 4.4 (3.1, 6.0) trillion. Net loss was largest in Maharashtra and smallest in Telangana. Sensitivity analyses confirmed that all ten states depicted losses in the case of a 3% discount rate, while five and seven states showed losses at 0% and 7% discount rates.

**Conclusion:** These are novel subnational estimates depicting annual net losses due to alcohol use in India with direct policy implications to disincentivize alcohol sales and raise taxes to an adequate level.

**Highlights:**

- The annual economic burden of alcohol use in India runs over six trillion rupees.
- Economic burden exceeds revenue from excise taxes noting net loss.
- Economic burden varies across states with Maharashtra having the largest burden.
- All of the top ten revenue-earning states depict net losses in primary analysis.

## 1 Introduction

Alcohol has been considered the most harmful drug passing heroin and crack cocaine (Nutt et al., 2010). Globally, alcohol use disorders are the most prevalent substance use disorders with over 100 million cases estimated annually (GBD 2016 Alcohol and Drug Use Collaborators, 2018). Alcohol use is also among the top risk factors associated with several morbidities. Over 99 million disability-adjusted life-years (DALYs) or 4% of the global disease burden can be attributed to alcohol use. Beyond health, there is also a known economic burden of alcohol use. Previous work has noted that the cost of alcohol use can be as high as 2.6% of a country’s gross domestic product (GDP) (Manthey et al., 2021). Similar evidence has been noted for India (Jyani et al., 2019).

Among other interventions, excise taxes on alcohol sales have been considered an effective means to reduce consumption and heavy drinking - thereby impacting morbidity and mortality (Guindon et al., 2022). However, the association between such health taxes and the expected health benefits is far more complex than is usually portrayed (see discussion ahead). Regardless, governments have often overlooked the public health costs of alcohol use and focused solely on revenue generation. Often, the popular Indian discourse has been that short-term economic gains from revenue generation take priority over long-term health concerns in developing economies (Zadey, 2020). In the current study, our first aim was to assess the economic burden of alcohol use at national and subnational levels. The second aim was to assess if this burden exceeded the revenue generation from excise taxes on alcohol sales.

## 2 Methods

We conducted a retrospective analysis for 2019 for India and its states. The analysis can be delineated into two parts. The first part focused on estimating the economic burden of alcohol use. For this, we used the disability-adjusted life-years (DALYs) for both sexes and all ages for all causes with alcohol use as the underlying risk factor from the Global Burden of Disease 2019 (GBD 2019 Diseases and Injuries Collaborators, 2020). Through this, we investigate the comprehensive disease burden that can be attributed to alcohol beyond alcohol use disorders. The economic burden associated with this disease burden is calculated using the value of life-year (VLY) or full-income approach. The Lancet Commission on Investing in Health notes that VLY accounts for the gains in GDP as well as those in life expectancy due to investments in healthcare interventions (Jamison et al., 2013). It also extends the human capital approach (that ties population health to a country’s GDP) to include those beyond the working or economically productive populations. For South Asia, the values of one life-year are considered to be equivalent to 2.8 and 1.2 times the GDP per capita at 3% and 7% discount rates, respectively. **Equation 1** describes how economic burden was calculated under different discount rates. Mean and 95% uncertainty interval values were extracted from GBD 2019 to propagate uncertainty from DALYs to VLYs.

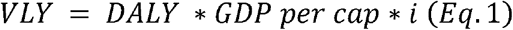

*where, i = 1 for 0% discount rate*

*= 2.8 for 3% discount rate*

*= 1.2 for 7% discount rate*

Data on national and state GDPs in Indian national rupees (INR) for the financial year 2019-20 was extracted from the Reserve Bank of India (RBI) handbook of statistics (Reserve Bank of India, 2020).

The second part of the analysis focused on estimating the net benefits (or losses) from the excise taxes on alcohol sales. Data on national and top ten states for excise tax revenues (in INR) for 2019-2020 was obtained from the news report citing RBI data (Kant, 2020). Annual net benefits or losses were calculated as per **Equation 2**. Three values of net losses/benefits were calculated using the three values for economic burden based on different discount rates. Uncertainty was propagated from the 95% intervals for VLY values.

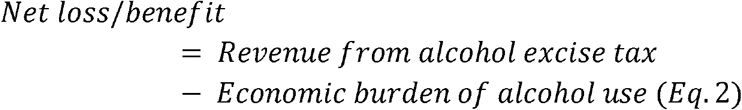

The VLY and net loss estimates based on the 3% discount rate are considered primary in the current analysis while those based on 0% and 7% discount rates are considered as sensitivity estimates. All monetary values are presented in 2019 INR. All data used and generated in the analysis is presented in **Supplementary Data**.

## 3 Results

### 3.1 Economic burden of alcohol use

Our primary analysis found that the economic burden of alcohol use in India was 6.24 (95%UI: 4.81, 7.77) trillion INR. Across states, it ranged from 7.31 (4.68, 10.40) billion INR for the northeastern state of Mizoram to 1.08 (0.76, 1.39) trillion INR for the western state of Maharashtra **(Figure 1A)**. The national and state-wise sensitivity analysis estimates are presented in **Supplementary Data**.

**Figure 1A:**
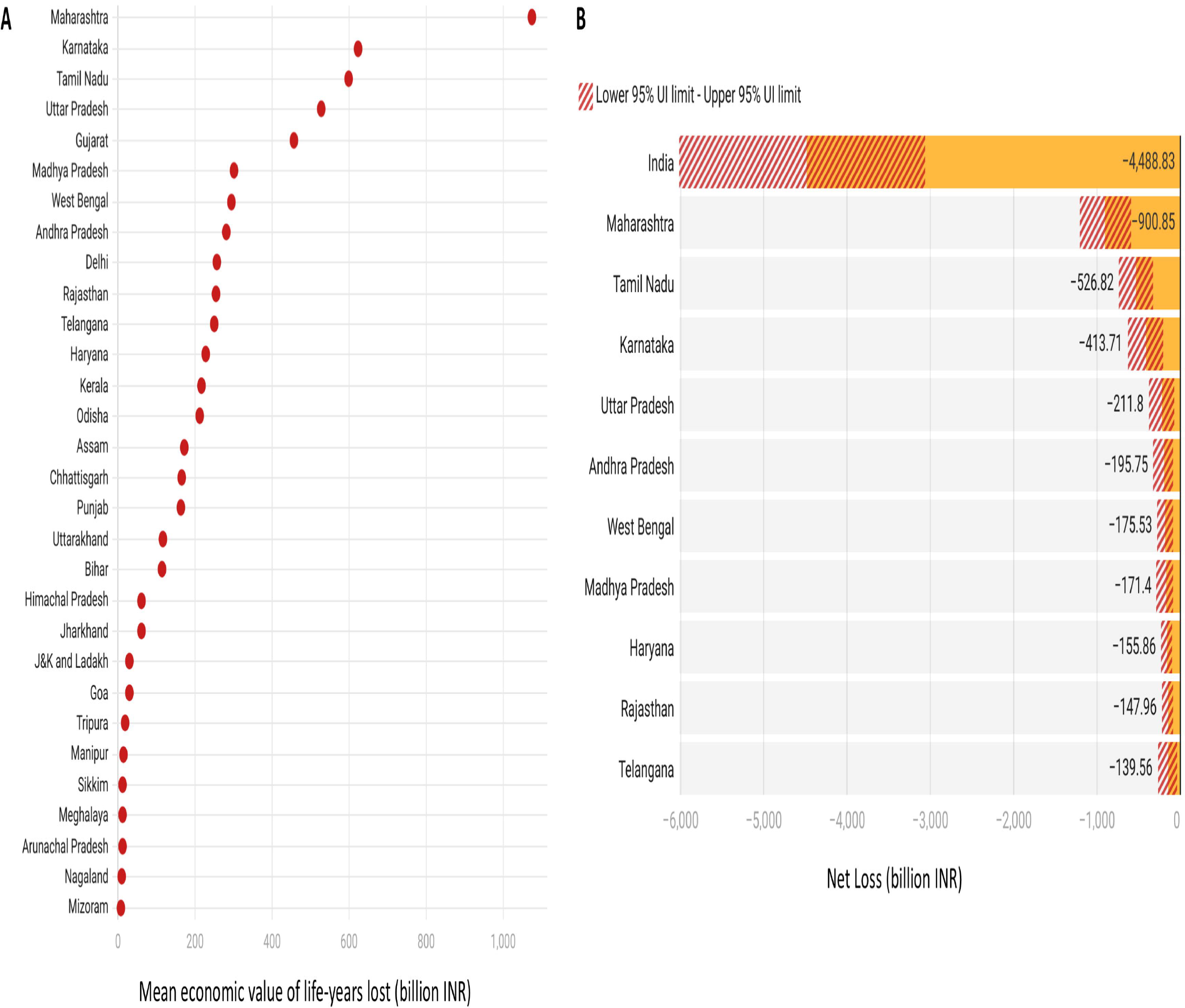
Economic burden (in billion INR) of alcohol use across 30 states sorted from largest to smallest burden. B. Net losses/benefits (in billion INR) relative to revenue generated from excise taxes on alcohol sales in the top ten revenue-earning states. 95% uncertainty intervals are presented by the shaded regions in B. Both A and B depict primary analysis conducted at a 3% discount rate.

### 3.2 Net losses due to alcohol use

In 2019, India collected 1.76 trillion INR in revenue from alcohol excise taxes. Hence, the net loss was 4.49 (3.06, 6.02) trillion INR. All the ten states with the topmost revenues from the alcohol excise taxes depicted net losses in the primary analysis **(Figure 1B)**. Maharashtra observed the highest loss of 0.90 (0.60, 1.21) trillion INR while Telangana noted the smallest net loss of 0.14 (0.04, 0.27) trillion INR. Sensitivity analysis also noted net loss nationally. Analysis using a 0% discount rate depicted a net loss in five of the ten states while that using a 7% discount rate had a net loss in seven states **(Figure 2A-B)**. The national and state-wise sensitivity analysis estimates are presented in **Supplementary Data**.

**Figure 2:**
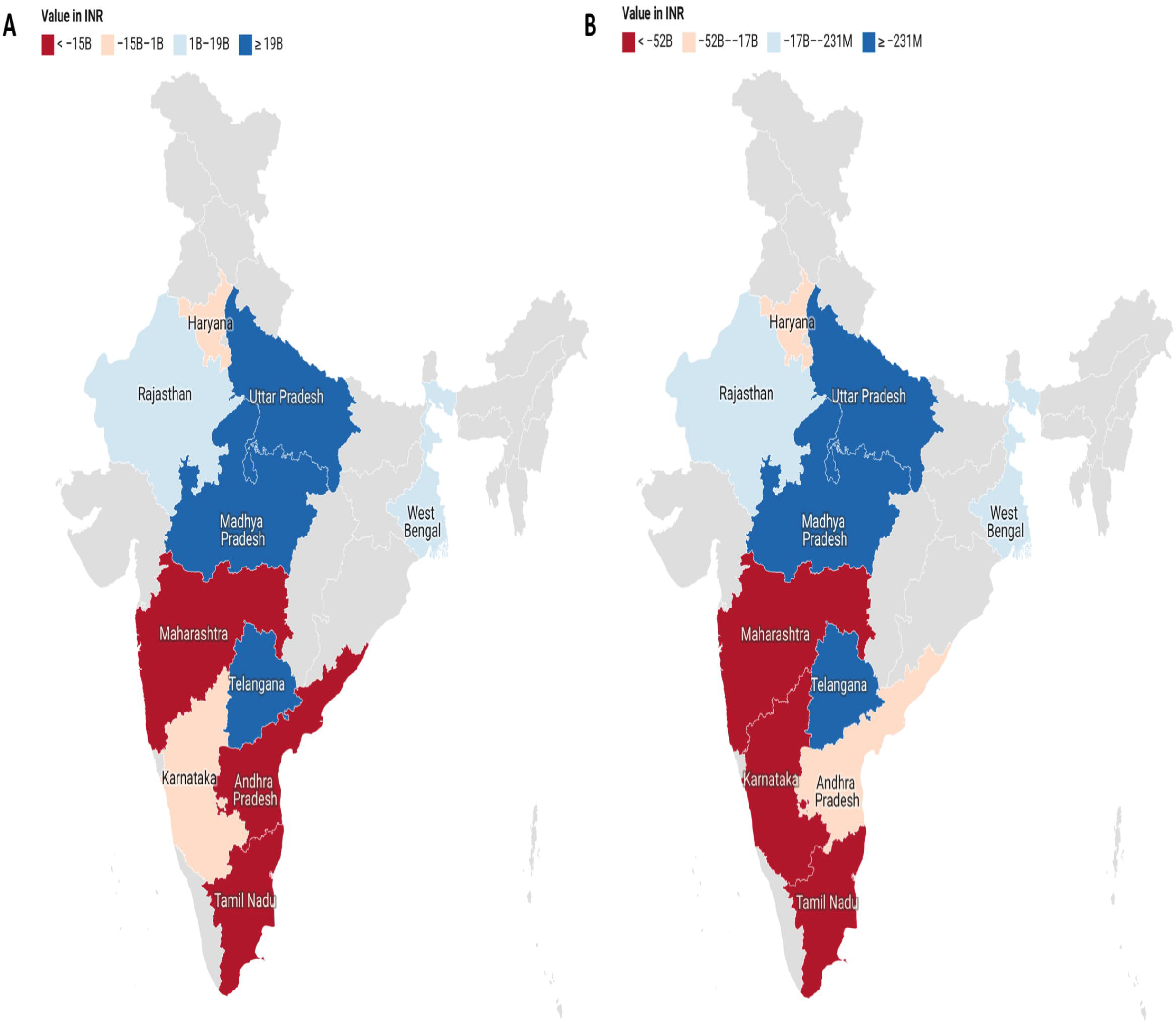
Net losses/benefits (in billion INR) relative to revenue generated from excise taxes on alcohol sales in the top ten revenue-earning states at A) 0% discount rate and B) 7% discount rate values.

## 4 Discussion

### 4.1 Summary of Findings

The current analysis used the value of life-year approach to estimate the annual economic burden of alcohol use in India and its 36 states/union territories for 2019. Additionally, we also found that this burden surpasses the revenue generated by excise taxes on alcohol resulting in net losses for the country and its top ten revenue-earning states. Sensitivity analyses depict that some states might not face net losses under differing discount rate assumptions. However, the primary and sensitivity analyses agree in most instances.

### 4.2 Literature Context

Literature on the economic burden of alcohol use is common among high-income countries. However, data on low- and lower-middle-income countries, including India, is limited. Previously, a technical brief from the World Health Organization summarized available literature to conclude that the economic burden of alcohol use could run up to 2.26% of the GDP annually (World Health Organization - Country Office for India & MSG Strategic Consulting, n.d.). However, this brief only used the 2013-14 costing data on healthcare, crime and law enforcement, road traffic injuries, and productivity losses. For comprehensive estimation, we used the ‘all cause’ category with alcohol use as a risk factor from the GBD 2019. More recently, a modeling study demonstrated that the accumulating economic burden due to alcohol use including the direct and indirect costs in the 2011 to 2050 period could be as high as 121 trillion INR (Jyani et al., 2019). Further, it also noted that the burden after adjusted for alcohol sales taxes will be about 98 trillion INR with a projected annual loss of 1.45% of the GDP to country. The current analysis agrees with the essence of the previous findings. We found the annual economic burden of alcohol use can run into trillions and is much above the revenue generated from alcohol excise taxes leading to net economic losses. The current analysis builds on previous work in two important ways. While modeling studies have depicted long-term losses, it is often argued that the government has the incentive of revenue generation in the short run. This analysis shows annual losses, countering that popular argument. Further, we provide subnational or state-level estimates of economic burden and net losses that were missing previously.

### 4.3 Policy Implications

From a public health perspective, the economic burden of alcohol use is much higher than the excise taxes collected from alcohol sales. This should encourage the state governments to consider the following. First, raise the excise taxes to a level that leads to some health gains in terms of reducing the incidence of alcohol consumption and heavy drinking. We do not intend to imply that the taxes should cover all losses. Hence, it is noteworthy, that our estimates for net losses should be treated as ‘upper bound’ estimates. Second, avoid using revenue generation as a way to discount harms due to alcohol use. Third, policymaking needs to reorient toward the notion that the short-term (annual) economic burden of alcohol use is immense. Since our analysis includes only cross-sectional data for 2019, it does not consider the lag between taxing alcohol consumption and it impact on reducing the economic burden associated with alcohol use. While that is a crucial assessment, it is not what we intend to investigate here. The current analysis rather argues that taxes should consider the past burden for better formulation ahead.

It is important to note that while alcohol sales taxes are widely supported, multiple design and implementation issues can prevent their desired impact (Wright et al., 2017). First, such taxes can be regressive putting a greater burden on lower socioeconomic sections than others. For India, this can exacerbate existing health and wealth disparities among rural and urban residents and people from different caste and religious groups. Second, taxes can encourage alcohol consumers to substitute taxed products with non-taxed alternatives. In India, such substitution is known to be common with country liquor - which may not always be safe. Third, the level of taxes should be high enough to prevent the rise in alcohol consumption. Our analysis notes that this isn’t the case in India. Finally, the revenue from such taxes should ideally be year-marked for health investments but that does not always happen. In Indian states, the taxing decisions and influenced by several factors and it is unclear whether the revenue generated from the alcohol excise taxes is invested back into public health. In India, previously, it has been shown that the price elasticity for demand, i.e., change in alcohol demand associated with a 1% increase in alcohol price, ranged from −0.14 for spirits to −0.46 for country liquor (Kumar, 2017). This points to only a modest reduction in consumption through price control. Similar issues have been pointed out in other low - and lower-middle-income countries, including those in Africa where taxes on alcohol sales may not always result in the desired health outcomes (Bird & Wallace, 2010).

### 4.4 Strengths and Limitations

The present study provides novel and recent subnational-level estimates for the economic burden of alcohol use and net losses compared to excise taxes on alcohol sales in India. To ensure the robustness of findings, sensitivity estimates at different discount rates are presented along with uncertainty propagation throughout different stages in the analysis. However, we acknowledge multiple limitations. First, this was a cross-sectional analysis involving only annual estimates without any projections for the future. Second, our analysis was macroeconomic in design and did not consider different components of the economic burden such as healthcare costs, indirect costs, productivity losses, etc. Third, we did not provide disaggregated estimates by sexes, age groups, rural/urban residence, etc. Fourth, we could not include smaller states and union territories in the economic burden analysis due to the lack of disease burden data on them. Fifth, for net benefits/losses analysis, we focused only on the national value and top ten revenue-earning states due to easy data availability, excluding other states. Regardless, the study’s findings have direct implications for policymaking and reorienting the narrative around the economic burden of alcohol use in the context of revenue generation. Future assessments should strengthen the methodological approach and validate these findings.

## Data Availability

All data produced in the present work are contained in the manuscript's Supplementary Material.

## 6 Declarations

### 6.1. Author Contributions

Siddhesh Zadey is the sole author of the manuscript and has contributed to Conceptualization, Methodology, Formal Analysis, Data Curation, Writing - Original Draft, Writing - Review & Editing, Supervision, and Project Administration.

## 6.3 Acknowledgements

None

## 6.5 Ethics approval and consent to participate

This manuscript uses publicly available aggregate-level data. Hence, ethics approval and consent obligations are not applicable.

## 6.6 Availability of data and materials

All data used and created in the manuscript is included in the Supplementary Material.

## 6.7 Declaration of Generative AI and AI-assisted technologies in the writing process

During the preparation of this work the author did not use any such tool. The author takes full responsibility for the content of the publication.

